# Mental Health Utilization in Children in the time of COVID-19

**DOI:** 10.1101/2021.08.11.21261712

**Authors:** Leah Coates, Rebecca Marshall, Kyle Johnson, Byron A. Foster

**Author notes:** **Corresponding author:** Byron A Foster, MD, MPH, 3181 SW Sam Jackson Park Rd., CDRC, Portland, OR 97239.

## Abstract

**Background:** In early 2020, coronavirus disease 2019 (COVID-19) was declared a public health emergency and a combination of lockdown and social distancing measures were put in place across the globe. Many children, adolescents and adults have experienced adverse mental health effects related to the pandemic and its impact on daily life, although the long-term impact on individuals and health systems is not well understood.

**Methods:** This cross-sectional study was based on data from 2018-2021 collected via medical records from our hospital. Admissions were transformed into time-series data, and models were generated to analyze changes in admission rates for mental health emergencies in 2020 and 2021 compared to previous years.

**Results:** Of 1906 inpatient encounters among 1543 unique patients seen by the Child and Adolescent Psychiatry Consultation-Liaison service, there was a decrease in overall admissions beginning in March 2020, coinciding with statewide lock down due to the COVID-19 pandemic. In April 2020, admissions were reduced 36% compared to average admissions from 2018-2019. By 2021, overall admissions were significantly higher than for the previous three years. Similarly, the count of suicide attempts was significantly higher in 2021 compared to previous years. The rate of patients admitted to inpatient facilities upon discharge was significantly higher during the COVID-19 pandemic period.

**Conclusion:** Admissions for mental health emergencies fluctuated during the period associated with the COVID-19 pandemic across an array of diagnoses. Increases in admissions and severity of mental health emergencies during COVID-19 may reflect a detrimental impact of the pandemic on the mental health of children, as well as unmet needs during this time.

## Background

The first laboratory-confirmed cases of coronavirus disease (COVID-19) were reported in the United States in January 2020 (1), and in subsequent months, measures to slow the spread of infection such as isolation and social distancing were widely implemented. Many of the measures implemented to prevent viral spread had significant impacts on daily life, including decreased healthcare access, social isolation, and disrupted routines, including school routines for children; they have been associated with adverse mental health effects (3). A growing body of evidence suggests that the COVID-19 pandemic has resulted in worsened anxiety and depression-related illness, reflective of fear of disease as well as lockdown and social distancing measures (3).

Children and adolescents may be more vulnerable to the psychosocial effects of a pandemic because they are in a critical period of development (4). National emergency department (ED) data from April through October 2020 reported increased proportions of children’s mental health visits among all pediatric visits, with overall increases in mental-health related visits compared to 2019. Recent data suggest a more marked increase of suicide attempts and suicidal thinking among 12-25 year old children in the first part of 2021 (5).This data underscores the negative impact of the COVID-19 pandemic on the mental health of children (6), possibly mediated through lack of access to routine mental health care, including school-based support (7) (8).

The principle aim of this project was to identify the impact of the COVID-19 pandemic on the frequency of mental health emergencies, using data from our hospital. The secondary aim was to examine the association between the COVID-19 pandemic and the severity of childhood mental health emergencies seen at our hospital. We hypothesize that during the COVID-19 pandemic time period, we will observe an increase in hospital admissions for childhood mental health services and an increase in the severity of mental health crises compared to the period prior to the pandemic.

## Methods

### Study Design and Participants

This is a descriptive study of patients seen by the child and adolescent psychiatry consult/liaison (CAP CL) service of our hospital, an academic teaching hospital in the Pacific Northwest, with 150 medical beds. Participants were eligible for inclusion if they were admitted between January 1, 2018, and April 30, 2021. There were no exclusion criteria. Participants were filtered according to whether their admission date occurred before or after the onset of the COVID-19 pandemic, defined as March 16, 2020, coinciding with state-wide school closures in the state. Outcome measures included frequency and severity of mental health emergencies seen at our hospital before and during the COVID-19 pandemic. This study was approved by our hospital’s institutional review board (IRB); because the dataset was large and the study was retrospective, informed consent was not obtained, and a waiver of consent approved by the IRB. This manuscript follows the Strengthening and Reporting of Observational Studies in Epidemiology (STROBE) reporting guideline for cross-sectional studies.

### Exposure of interest

The exposure of interest was the COVID-19 time-period, which was measured using admissions dates from medical records. Admissions were grouped by month and year to compare admissions that occurred during the COVID-19 pandemic to admissions that occurred prior to the pandemic. 2018-2020 data was available for all 12 calendar months, and 2021 data was available for the months of January through April.

#### Covariates of interest

The covariates of interest included age, gender, and insurance status. All covariates were retrieved from medical records. Age was reported in the medical charts and cross-referenced by calculating the amount of time between birthdate and hospital admission date. Gender was self-reported and categorized as male, female, transgender male, transgender female, or other. Insurance coverage was reported by insurance company, and categorized as commercial, government, other, or none.

### Outcomes of interest

The primary outcome of interest was the frequency of mental health crises during the COVID-19 pandemic compared to the time-period prior. Frequency was measured by the number of monthly and annual admissions over time, derived from daily hospital admissions records from our hospital. The secondary outcome of interest was the severity of mental health crises during the COVID-19 pandemic. We examined severity using the outcomes of suicidality, the referral unit, and the disposition for each admission (**Table 1**). Suicidality was measured as a four-level categorical variable and cases were considered severe if they were classified as suicidal. Referral unit was measured as a three-level categorical variable and indicated which department entered the initial referral to psychiatry, with cases seen in the PICU classified as severe. Disposition was measured as a four-level categorical variable indicating the type of care facility the patient was discharged to. Patients admitted to inpatient facilities were flagged as severe. The count of monthly and annual admissions classified as higher severity in the context of the variables were calculated and analyzed for changes in frequency over time and used as a proxy for severity of mental health crises. Hospital length of stay (LOS) was measured in days from the date of admission to the date of discharge and monthly mean or median LOS was visualized over time. Longer hospital LOS was also used as a proxy for severity.

**Table 1.** Description of outcome variables used to measure severity of mental health crises.

| Variable | Measured (levels) | Admissions (%) | Operationalized (*severity indicating level of variable) |
| --- | --- | --- | --- |
| Suicidality | Attempt | 628 (32.9%) | *Admission classified as a suicide attempt – level of variable flagged as severe |
|  | Plan/Intent | 73 (3.8%) | Admission classified as suicide plan or intent |
|  | Ideation | 292 (15.3%) | Admission classified as suicidal ideation |
|  | None | 913 (47.9%) | Admission classified as no suicidality |
| Referral unit | PICU | 312 (16.4%) | *Patient seen in the PICU – level of referral unit variable flagged as severe |
|  | ED | 954 (50.1%) | Patient seen in the ED |
|  | Hospital | 640 (33.6%) | Patient seen in the hospital |
| Disposition | Inpatient | 407 (21.4%) | *Patient admitted to inpatient facility upon discharge – level of disposition variable flagged as severe |
|  | Subacute | 148 (7.8%) | Patient admitted to subacute facility upon discharge |
|  | Residential | 28 (1.5%) | Patient admitted to residential facility upon discharge |
|  | Outpatient | 1321 (69.3%) | Patient admitted to outpatient facility upon discharge |

### Statistical Analysis

Encounter-level clinical and administrative data were pulled from hospital medical records and included admission dates, gender, age, insurance status, county of residence, hospital length of stay, suicidality, and disposition. We examined daily admissions to the hospital, ED, and PICU for mental health emergencies from January 2018 to April 2021. Admissions were sorted by year and described with summary statistics (**Table 2**). Admission counts were grouped by month and year, transformed into time series data, and displayed graphically (**Figure 1**). Poisson regression was used to examine changes in rate of hospital admissions with respect to the severity-indicating variables, both by year, and with respect to the COVID-19 pandemic. Admissions numbers were regressed by year, as well as the period following the onset of school closures (March 16, 2020). To account for changes in overall admissions, counts of cases with respect to specific severity-indicating variables were also displayed as proportions over total admissions numbers seen by the psychiatry department. Monthly trends in admission rates for the years 2018 and 2019 were displayed with locally estimated scatterplot smoothing. Monthly data from years 2020-2021 were displayed against these models for visual comparison (**Figure 1**). All statistical analyses were performed in R Studio version 1.2.5001 with the following packages: Methods, dplyr, lubridate, tidyr, ggplot2, and tidyverse. The threshold value for statistical significance was set at .05.

**Table 2.** Demographic characteristics stratified by pre-pandemic vs. pandemic time period

|  | Pre-Pandemic | Pandemic | Overall | P-Value |
| --- | --- | --- | --- | --- |
| **N** | 1255 | 651 | 1906 | 0.16 |
| **Age (y)** |  |  |  |  |
| **Mean (SD)** | 13.9 (2.74) | 13.8 (2.71) | 13.9 (2.73) | 0.23 |
| **Median [Min, Max]** | 15.0 [2.0, 19.0] | 14.0 [3.0, 18.0] | 14.0 [2.0, 19.0] | 0.10 |
| **Gender** |  |  |  |  |
| **Female (%)** | 750 (59.8%) | 401 (61.6%) | 1151 (60.4%) | 0.46 |
| **Male (%)** | 458 (36.5%) | 209 (32.1%) | 667 (35.0%) | 0.06 |
| **Trans Male (%)** | 44 (3.5%) | 17 (2.6%) | 61 (3.2%) | 0.34 |
| **Trans Female (%)** | 2 (0.2%) | 14 (2.2%) | 16 (0.8%) | **<0.001** |
| **Other** | 1 (0.1%) | 10 (1.6%) | 11 (0.6%) | **<0.001** |
| **Hospital Length of Stay** |  |  |  |  |
| **Mean (SD)** | 3.91 (5.90) | 4.56 (6.72) | 4.13 (6.2) | **0.03** |
| **Missing** | 8 (0.6%) | 0 (0%) | 8 (0.4%) | - |
| **Insurance** |  |  |  |  |
| **Commercial (%)** | 580 (46.2%) | 314 (48.2%) | 894 (46.9%) | 0.44 |
| **Government (%)** | 627 (50.0%) | 322 (49.5%) | 949 (49.8%) | 0.86 |
| **Other (%)** | 24 (1.9%) | 6 (0.9%) | 30 (1.6%) | 0.15 |
| **None (%)** | 23 (1.8%) | 9 (1.4%) | 32 (1.7%) | 0.59 |
| **Missing (%)** | 1 (0.1%) | 0 (0%) | 1 (0.1%) | - |

**Bold:** p<0.05

**Figure 1.**
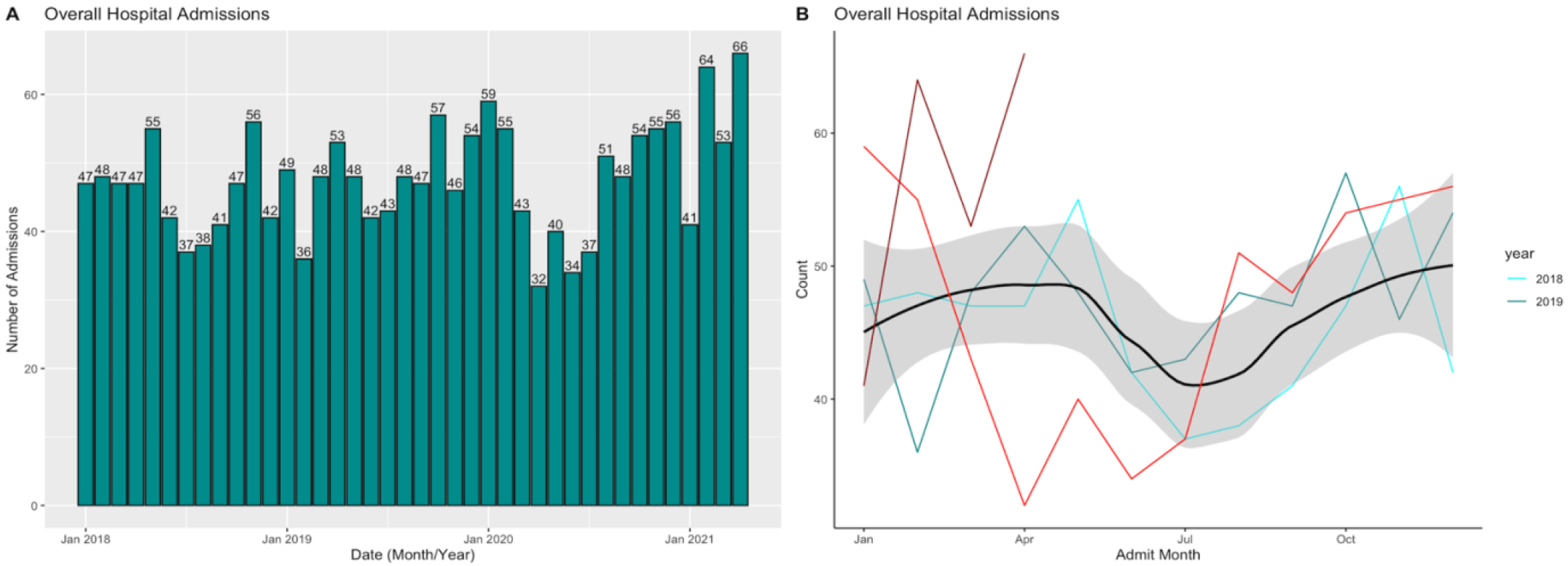
Monthly psychiatric admissions from January 1, 2018 - April 30, 2021. Monthly overall admission rates for the psychiatry service displayed as A. Bar graph displaying raw monthly admissions numbers and B. Loess plot with a smoothed trendline for monthly admissions for years 2018-2019 with 2020 admissions overlaid in red and 2021 admissions overlaid in dark red.

## Results

### Demographic Characteristics

There were 1,906 encounters among 1,543 unique patients between January 1, 2018, and April 30, 2021. Approximately 32.7% of the encounters occurred between March 16, 2020, and April 30, 2021. 14.9% of the admissions that occurred during this period were repeat admissions. As is shown in **table 2**, the cohort had a median (interquartile range [IQR]) age of 14 (13-16) years and included 1151 (60.4%) individuals who identified as female, 667 (35.0%) individuals who identified as male, 61 (3.2%) individuals who identified as trans-male, 16 (0.8%) individuals who identified as trans-female, and 11 (0.6%) individuals who identified as other. A total of 949 (49.8%) patients had government insurance, and the median (IQR) length of stay for the 1,906 encounters was 2 (1-5) days.

### Seasonal Patterns in Mental Health Emergencies

Overall admissions to our hospital between 2018-2021 are displayed in **Figure 1**. Differences in admissions for winter months in relation to summer months were calculated by comparing the number of admissions occurring in each season respectively for years 2018-2020. There was an overall greater number of total admissions in winter months compared with summer months, however there was not a significant difference between the two seasons (mean [SD], 51.7 [6.4] vs 39.0 [3.5]; P=0.12). Similarly, there was not a greater number of mental health emergencies classified as suicide attempts occurring in the winter months during these years compared with the summer months (mean [SD], 14.7 [3.2] vs 11.3 [1.5]; P=0.41).

### Changes in Admissions Patterns During the COVID-19 Pandemic

Deviations from previous admissions patterns for the years 2020-2021 are displayed in **Figure 2** (2020 shown in red, 2021 shown in dark red). During January – February of 2020, the overall number of children’s mental health related hospital visits was elevated compared to the average from the previous two years by approximately 26.7%. During March and April of 2020, mental health related hospital visits fell below counts from previous years by approximately 23.1% coinciding with widespread shelter-in-place orders. Hospital visits remained below the average for previous years until July when they began to steadily increase, reaching previous averages in August, and continuing to climb through the fall months. As of February 2021, overall admissions were higher than averages from the previous three years (2018-2020) by 38.1%. 2021 admissions declined again in March, but by April they were higher than previous year’s averages by 50%.

**Figure 2.**
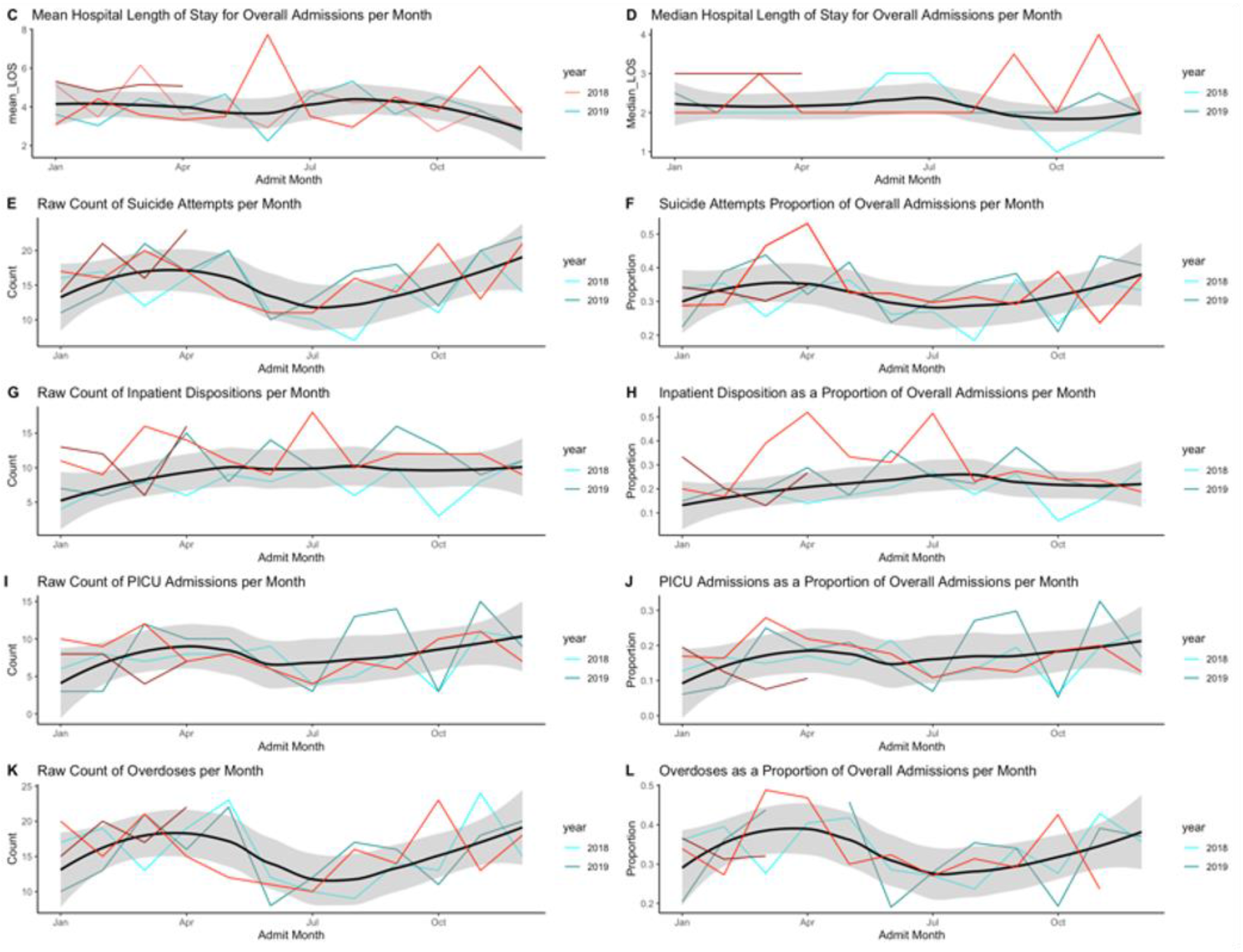
Seasonal admissions patterns for selected severity indicating variables, January 1, 2018 - April 30, 2021. Seasonal patterns for: A. mean hospital length of stay, B. median hospital length of stay, C. raw count of suicide attempts, D. suicide attempts proportion of overall admissions, E. raw count of inpatient dispositions, F. inpatient dispositions as a proportion of overall admissions, G. raw count of PICU admissions, H. PICU admissions as a proportion of overall admissions, I. raw count of overdoses, J. Overdoses as a proportion of overall admissions. Smoothed trendline for 2018-2019 seasonal patterns displayed with 2020 seasonal patterns overlaid in red and 2021 patterns overlaid in dark red.

Overall admissions were not significantly increased during the total period of the COVID-19 pandemic compared to prior, but admissions were significantly higher for the year 2021 compared to the previous three years (p=0.03). Similarly, the count of admissions classified as suicide attempts was found to be significantly elevated during the year 2021 compared to previous years (p<0.001), but not during the COVID-19 pandemic overall. During the COVID-19 pandemic, the number of patients admitted to inpatient facilities upon discharge was significantly higher compared to the time prior (p<0.001). Additionally, the number of patients discharged to inpatient facilities was also significantly higher during years 2019-2021 compared to 2018 (p<0.001). The mean hospital length of stay for admissions prior to the COVID-19 pandemic was 3.9 days compared to 4.6 days after the onset of the pandemic (p=0.03), indicating a significantly longer length of stay for patients seen during this time. (**Table 3**).

**Table 3.** Comparison of admissions before/During the COVID-19 pandemic and from 2018-2021.

|  |  | IRR | 95% CI |
| --- | --- | --- | --- |
| Overall admissions |  |  |  |
| COVID-19 | No | Reference |  |
|  | Yes | 0.93 | 0.76, 1.14 |
| Year | 2018 | Reference |  |
|  | 2019 | 0.96 | 0.80, 1.15 |
|  | 2020 | 0.95 | 0.80, 1.13 |
|  | 2021 | **1.23** | **1.02, 1.48** |
| Suicide attempts |  |  |  |
| COVID-19 | No | Reference |  |
|  | Yes | 1.05 | 0.88, 1.24 |
| Year | 2018 | Reference |  |
|  | 2019 | 1.15 | 0.94, 1.41 |
|  | 2020 | 1.04 | 0.85, 1.27 |
|  | 2021 | **1.31** | **1.02, 1.68** |
| Inpatient admissions |  |  |  |
| COVID-19 | No | Reference |  |
|  | Yes | **1.26** | **1.04, 1.53** |
| Year | 2018 | Reference |  |
|  | 2019 | **1.41** | **1.11, 1.80** |
|  | 2020 | **1.47** | **1.18, 1.83** |
|  | 2021 | **1.57** | **1.10, 2.22** |
| PICU admissions |  |  |  |
| COVID-19 | No | Reference |  |
|  | Yes | 0.90 | 0.72, 1.11 |
| Year | 2018 | Reference |  |
|  | 2019 | 1.16 | 0.97, 1.39 |
|  | 2020 | 1.03 | 0.86, 1.23 |
|  | 2021 | 0.93 | 0.77, 1.12 |
| Overdoses |  |  |  |
| COVID-19 | No | Reference |  |
|  | Yes | 1.00 | 0.84, 1.19 |
| Year | 2018 | Reference |  |
|  | 2019 | 0.98 | 0.78, 1.23 |
|  | 2020 | 0.92 | 0.74, 1.15 |
|  | 2021 | 1.18 | 0.95, 1.47 |

IRR: Incident-rate ratios, **Bold:** p<0.05.

## Discussion

This study examined the impact of COVID-19 on mental health admissions for children and adolescents at a large children’s hospital using monthly data on the total number and severity of mental health emergencies seen before and during the COVID-19 pandemic. In 2020 during the months of March and April, we observed a substantial decline in the overall reported number of children’s mental health emergencies, coinciding with the widespread implementation of COVID-19 mitigation measures. These findings are consistent with previous studies which have found that overall reported numbers of mental health related ED visits for children declined between 65-70% between March 29-April 25 2020, compared to the same span of months from 2019 (6) (add citation: Sheridan 2020). In addition, along with the decrease in overall children’s mental health-related ED visits, the proportion of all ED visits for children’s mental health-related concerns increased compared to rates from 2019 (6). Describing both the numbers and proportions of mental health admissions provides additional context for the severity of mental health emergencies experienced during a time when non-emergent hospital visits were discouraged.

In this study, the proportion of admissions classified as suicide attempts was elevated in March-April 2020 compared to the same period from 2018-2019; however, the rate of pediatric admissions for suicide attempts was not initially higher following the onset of the COVID-19 pandemic. This finding is consistent with a previous study which examined positive results on suicide-risk screens routinely administered in a pediatric ED (9). The findings demonstrated that positive screens for recent suicide attempts were higher in February, March, April and July 2020 compared to the same months from 2019, although not overall significantly increased compared to previous years (9). However, we did observe a substantially higher rate in the first few months of data from 2021, which is consistent with a recent national report of suicidality among adolescents in the same period (5). It is important to note that the overall number of clinical encounters was substantially reduced during the initial months of the COVID-19 pandemic, therefore direct comparison of rates across months and or years should be made conservatively.

We observed a significantly greater number of admissions for patients who identify as transgender during the pandemic as compared to the period prior (p<0.001). This finding suggests this population may need additional support during the ongoing pandemic, as they may be at increased risk for mental health challenges. (10).

Changes to children’s lives due to the COVID-19 pandemic may produce positive and negative effects on mental health (11). For many, the COVID-19 pandemic has resulted in family economic hardship, limited access to services, and limited social contact, among other stressors; these may contribute to mental health challenges for children and parents (4) (11) (12). Many children receive mental health support through community agencies including schools (8), so the observed increase in the proportion of hospital visits coinciding with widespread school closures may reflect decreased access to previously available mental health support services (6). The data described here suggest an initial drop in mental health needs that may have been an artifact of an overall decrease in utilization of any kind of healthcare. The sustained elevation in suicidality admissions in 2021, consistent with the CDC’s recent report, may suggest a cumulative effect of isolation on adolescents.

There are many questions regarding the long-term effects of the pandemic on children and families, some of which may be positive.

## Limitations

The results of this study must be considered carefully in the context of the study limitations. The present study utilizes data from a single hospital, potentially limiting the external validity of findings. Approximately 15% of patient encounters for mental health emergencies that occurred during the time of the COVID-19 pandemic were repeat encounters, seen at least once prior since January 1, 2018. This may indicate that patients seen in the hospital, ED, or PICU constitute a high-risk population and therefore rates of suicide attempts reported in this study may not be reflective of true rates within the population. We are unable to draw causal inference due to the cross-sectional nature of the study, and the potential influence of additional social and historical-sociopolitical factors occurring during the studied period. Future studies may consider examining the long-term impact of the COVID-19 pandemic on childhood mental health and further study is warranted as additional data becomes available.

## Conclusions

This study provided insight on children’s mental health in the context of the COVID-19 pandemic and the associated burden on the healthcare system. Children’s mental health status can have both short and long-term implications for their overall health and wellbeing (5). The collection and interpretation of children’s mental health data is vital for monitoring the impact of the COVID-19 pandemic on mental health status. Visualizing changes in typical patient admission patterns during unique circumstances such as the COVID-19 pandemic is beneficial for planning and creating appropriate response strategies and policies. Ensuring availability and access to mental health support services for children is vital to promote wellbeing in general, especially during the COVID-19 response and recovery periods.

## Data Availability

Due to ethical concerns, supporting data cannot be made openly available.

